# BONEcheck: a digital tool for personalized bone health assessment

**DOI:** 10.1101/2023.05.10.23289825

**Authors:** Dinh Tan Nguyen, Thao P. Ho-Le, Liem Pham, Vinh P. Ho-Van, Tien Dat Hoang, Thach S. Tran, Steve Frost, Tuan V. Nguyen

## Abstract

**Background and Aim:** Osteoporotic fracture is a significant public health burden associated with increased mortality risk and substantial healthcare costs. Accurate and early identification of high-risk individuals and mitigation of their risks is a core part of the treatment and prevention of fractures. We aimed to introduce a digital tool called ’BONEcheck’ for personalized bone health assessment.

**Methods:** The development of BONEcheck primarily utilized data from the prospective population-based Dubbo Osteoporosis Epidemiology Study and the Danish Nationwide Registry. BONEcheck has 3 modules: input data, risk estimates, and risk context. Input variables include age, gender, prior fracture, fall incidence, bone mineral density (BMD), comorbidities, and genetic variants associated with BMD. By utilizing published methodologies, BONEcheck generates output related to the likelihood of fracture and its associated outcomes. The vocabulary utilized to convey risk estimation and management is tailored to individuals with a reading proficiency at level 8 or above.

**Results:** The tool is designed for men and women aged 50 years and older who either have or have not sustained a fracture. Based on the input variables, BONEcheck estimates the probability of any fragility and hip fracture within 5 years, skeletal age, subsequent fracture, genetic risk score, and recommended interval for repeating BMD. The probability of fracture is shown in both numeric and human icon array formats. The risk is also presented in the context of treatment and management options based on Australian guidelines. Skeletal age was estimated as the sum of chronological age and years of life lost due to a fracture or exposure to risk factors that elevate mortality risk. In its entirety, BONEcheck is a system of algorithms translated into a single platform for personalized osteoporosis and fracture risk assessment.

**Conclusions:** BONEcheck is a new system of algorithms that aims to offer not only fracture risk probability but also contextualize the efficacy of anti-fracture measures concerning the survival benefits. The tool can enable doctors and patients to engage in well-informed discussions and make decisions based on the patient’s risk profile. Public access to BONEcheck is available via https://bonecheck.org and in Apple Store (iOS) and Google Play (Android).

## Introduction

Osteoporosis is a skeletal condition characterized by reduced bone mass and deteriorated bone microstructure, leading to an increased risk of fracture. Globally, osteoporosis affects over 200 million people aged 50 years and older [1], with women being more susceptible than men. In addition, there are 178 million fractures, including 14.2 million hip fractures which is the most severe manifestation of osteoporosis [2]. An existing fracture is associated with an increased risk of further fractures [3] and an increased risk of premature mortality [4]. With the ongoing aging population worldwide, it is expected that the burden and consequences of fractures will be more pronounced in the future.

A significant proportion of osteoporotic fractures and fracture-associated deaths is preventable by either taking a treatment or preventive measures. However, at present, there is a crisis of osteoporosis management in which most patients with a fracture are not treated. Moreover, even among those on treatment, adherence has been poor, with many patients opting out of the treatment program [5]. Therefore, a research priority in osteoporosis is to identify high-risk individuals for treatment and prevention. Over the past two decades, fracture risk assessment tools such as the Garvan Fracture Risk Calculator [6] and FRAX [7] have been developed and implemented in clinical practice. These tools map clinical risk factors to the probability of fractures over 5-year or 10-year for an individual. The implementation of fracture risk calculators represents a significant advance in the field.

Despite the advance, existing fracture risk calculators have a number of limitations in terms of form and communication. First, they use probability as a metric of risk, which is not readily understood by laypeople and doctors alike, especially when the probability is not presented in the context of the treatment [8]. Second, the presentation of risk is in a purely numerical format which is known to be less effective than a frequency format [9]. Third, existing risk calculators do not assess the risk of refracture and mortality, which are highly relevant to an individual. The lack of post-fracture mortality assessment might have led to the underappreciation of osteoporosis as a serious disease. These major limitations call for a more effective and innovative risk assessment tool.

In this paper, we describe the development of a digital fracture risk assessment tool that addresses the above limitations. The new tool is called ’BONEcheck’ and is available free worldwide. It is anticipated that the tool will help facilitate doctor-patient communication about fracture risk, its survival consequences, and interventional options.

## Materials and Methods

BONEcheck includes 3 modules: input data, risk estimates, and risk interpretation/context. The input data are determined from previous studies that have identified relevant and independent risk factors for fracture. The output information is designed from patients’ perspectives and presented in a format that is meaningful to patients. This output is contextualized in relation to treatment and preventive measures. The language used in the interpretation of risk estimates and risk management is written for individuals whose reading level is at or above 8.

### Input data

BONEcheck uses a range of variables that capture the uniqueness of the risk profile for an individual. These variables include anthropometric data (e.g., age, gender, height, weight), lifestyle factors (e.g., smoking habit, alcohol consumption), and bone-related data such as femoral neck bone mineral density (BMD) and a personal history of fracture. We chose to use femoral neck BMD rather than lumbar spine BMD because the former has been shown to be less prone to artefactual errors due to degenerative changes. BMD can be entered either in absolute values (gram per cm^2^) or T-score, which is the number of standards deviation from the peak BMD was taken as aged between 20 and 30 years. Individuals with a T-score equal to or less than −2.5 is diagnosed to have ’osteoporosis’. The computation, however, uses actual continuous BMD measurement, not T-scores. The personal history of fracture is entered as the number of prior fractures, not a binary value of ’yes’ or ’no’.

In addition, BONEcheck also requires input data pertaining to fall history, existing comorbidities, and a genetic profile. The number of falls over the previous 12 months is used for estimating the risk of fracture. Existing comorbidities include a list of 11 chronic conditions (see **Table 1**), with each condition being entered as ’yes’ or ’no’. Based on the self-reported comorbidities, BONEcheck calculates the Charlson Comorbidity Index (CCI) [10], which is used as a risk factor for the estimation of post-fracture mortality and skeletal age.

**Table 1:**
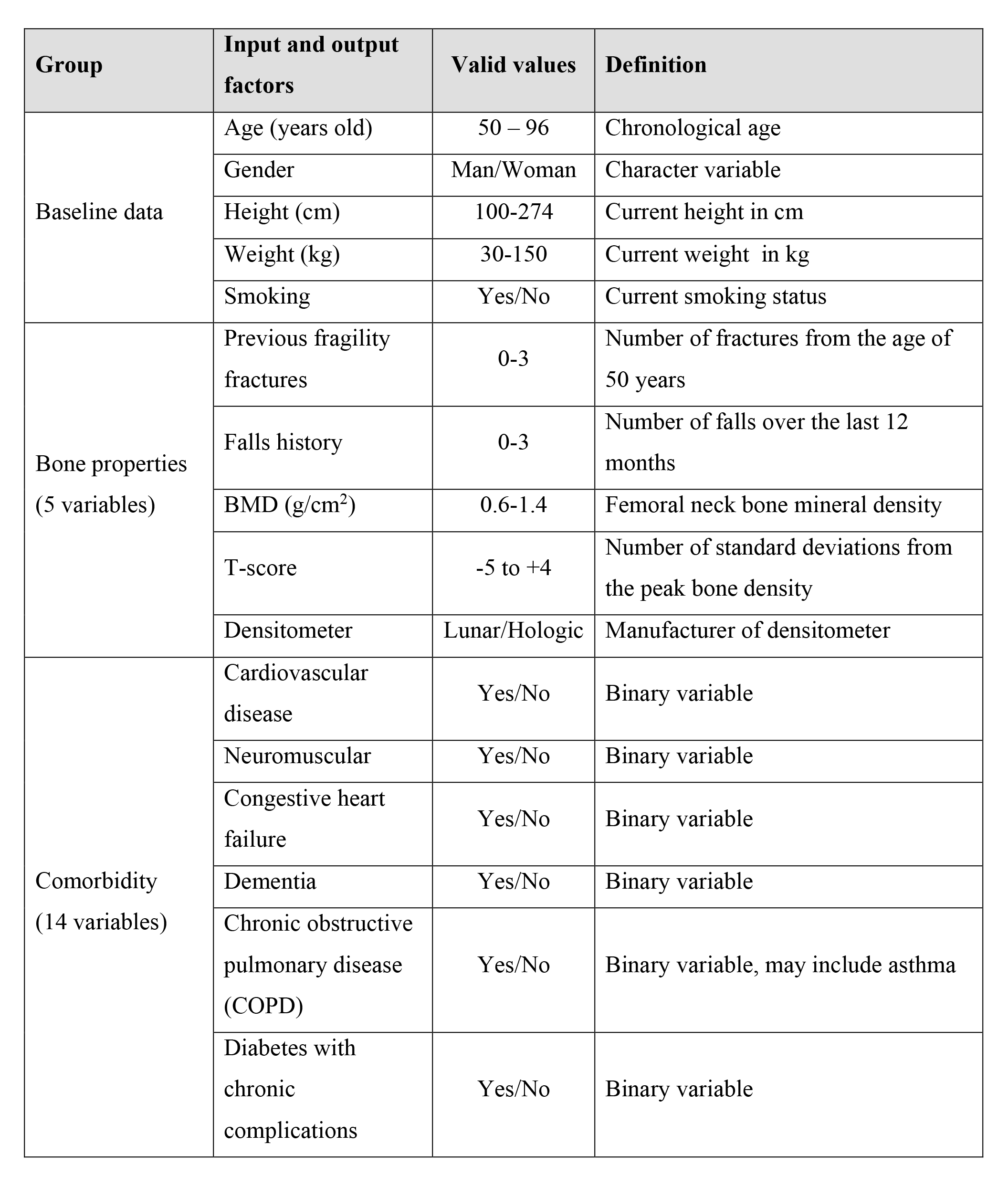
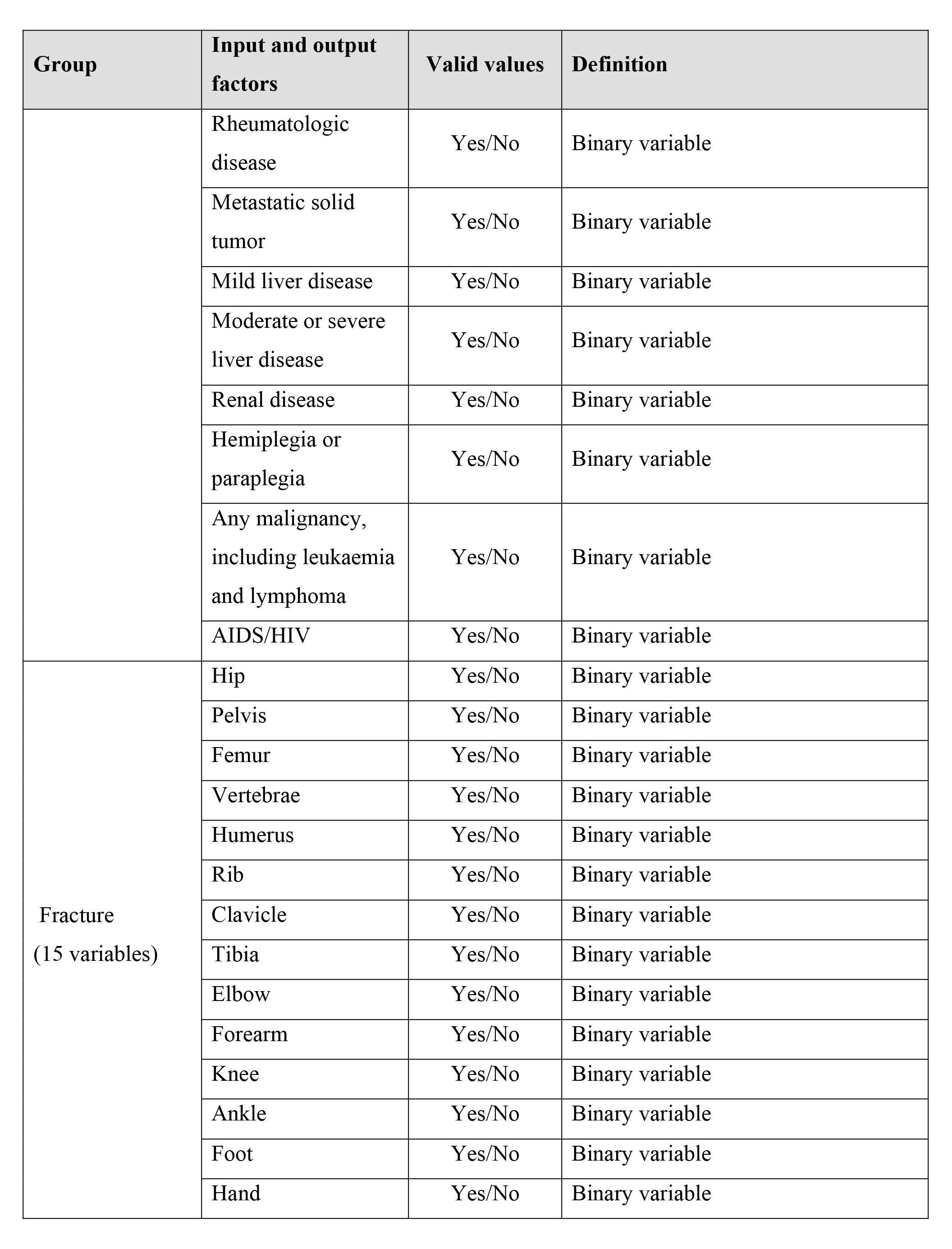
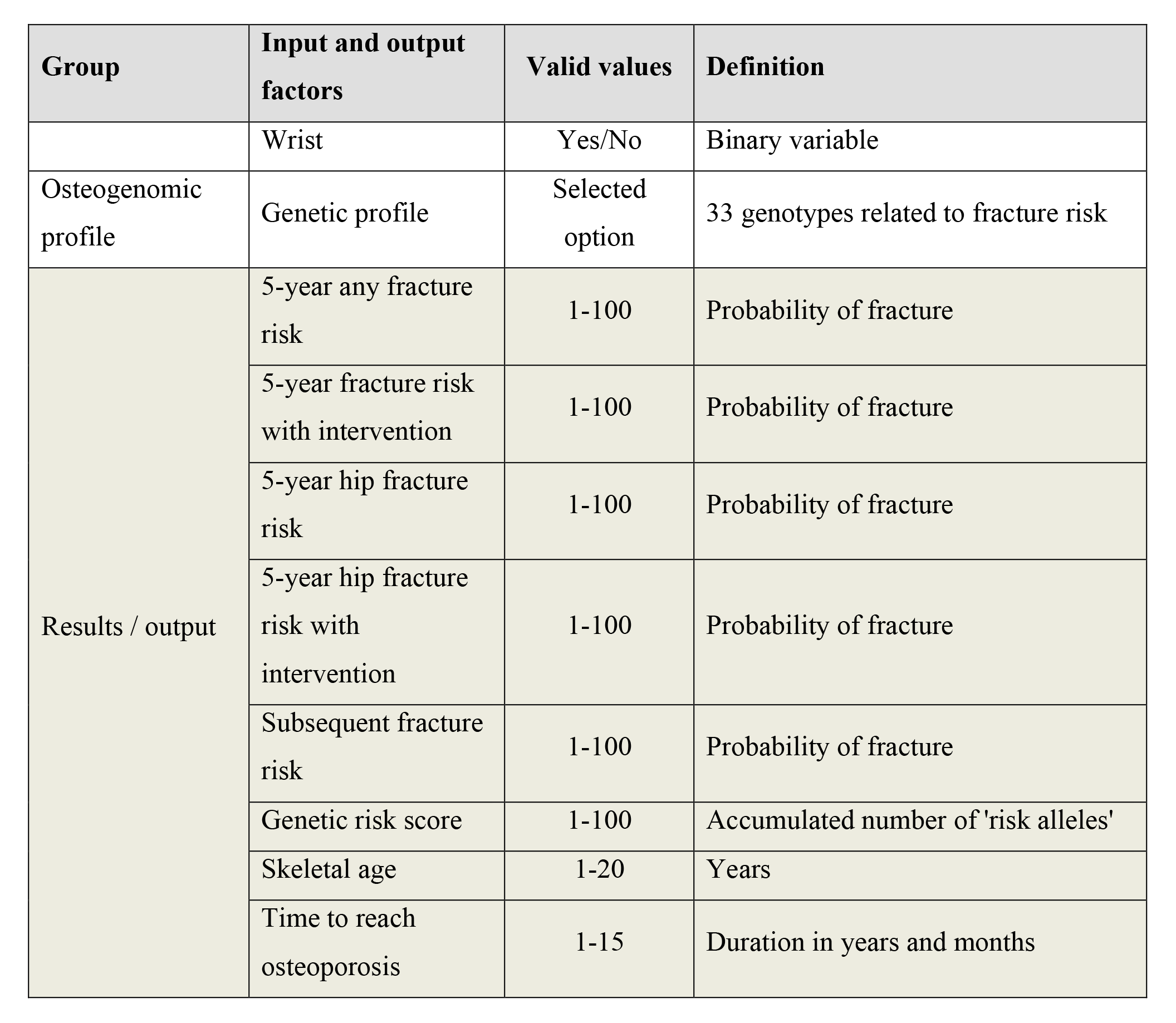
Input variables and output information in BONEcheck.

| **Group** | **Input and output factors** | **Valid values** | **Definition** |
| --- | --- | --- | --- |
| Baseline data | Age (years old) | 50 – 96 | Chronological age |
|  | Gender | Man/Woman | Character variable |
|  | Height (cm) | 100-274 | Current height in cm |
|  | Weight (kg) | 30-150 | Current weight in kg |
|  | Smoking | Yes/No | Current smoking status |
| Bone properties<br>(5 variables) | Previous fragility fractures | 0-3 | Number of fractures from the age of 50 years |
|  | Falls history | 0-3 | Number of falls over the last 12 months |
|  | BMD (g/cm<sup>2</sup>) | 0.6-1.4 | Femoral neck bone mineral density |
|  | T-score | -5 to +4 | Number of standard deviations from the peak bone density |
|  | Densitometer | Lunar/Hologic | Manufacturer of densitometer |
| Comorbidity<br>(14 variables) | Cardiovascular disease | Yes/No | Binary variable |
|  | Neuromuscular | Yes/No | Binary variable |
|  | Congestive heart failure | Yes/No | Binary variable |
|  | Dementia | Yes/No | Binary variable |
|  | Chronic obstructive pulmonary disease (COPD) | Yes/No | Binary variable, may include asthma |
|  | Diabetes with chronic complications | Yes/No | Binary variable |

| Group | Input and output factors | Valid values | Definition |
| --- | --- | --- | --- |
|  | Rheumatologic disease | Yes/No | Binary variable |
|  | Metastatic solid tumor | Yes/No | Binary variable |
|  | Mild liver disease | Yes/No | Binary variable |
|  | Moderate or severe liver disease | Yes/No | Binary variable |
|  | Renal disease | Yes/No | Binary variable |
|  | Hemiplegia or paraplegia | Yes/No | Binary variable |
|  | Any malignancy, including leukaemia and lymphoma | Yes/No | Binary variable |
|  | AIDS/HIV | Yes/No | Binary variable |
| Fracture<br>(15 variables) | Hip | Yes/No | Binary variable |
|  | Pelvis | Yes/No | Binary variable |
|  | Femur | Yes/No | Binary variable |
|  | Vertebrae | Yes/No | Binary variable |
|  | Humerus | Yes/No | Binary variable |
|  | Rib | Yes/No | Binary variable |
|  | Clavicle | Yes/No | Binary variable |
|  | Tibia | Yes/No | Binary variable |
|  | Elbow | Yes/No | Binary variable |
|  | Forearm | Yes/No | Binary variable |
|  | Knee | Yes/No | Binary variable |
|  | Ankle | Yes/No | Binary variable |
|  | Foot | Yes/No | Binary variable |
|  | Hand | Yes/No | Binary variable |
|  | Wrist | Yes/No | Binary variable |
| Osteogenomic profile | Genetic profile | Selected option | 33 genotypes related to fracture risk |
| Results / output | 5-year any fracture risk | 1-100 | Probability of fracture |
|  | 5-year fracture risk with intervention | 1-100 | Probability of fracture |
|  | 5-year hip fracture risk | 1-100 | Probability of fracture |
|  | 5-year hip fracture risk with intervention | 1-100 | Probability of fracture |
|  | Subsequent fracture risk | 1-100 | Probability of fracture |
|  | Genetic risk score | 1-100 | Accumulated number of 'risk alleles' |
|  | Skeletal age | 1-20 | Years |
|  | Time to reach osteoporosis | 1-15 | Duration in years and months |

The genetic profile included 34 genetic variants that have been shown to be associated with BMD in a genome-wide association study [11]. Each genetic variant is inputted as the number of minor alleles. Based on the data, an ’Osteogenomic Profile’ for each individual is generated as the weighted sum of the number of minor alleles across variants, with the weights being the published regression coefficient associated with each minor allele [12]. This Osteogenomic Profile, which has been shown to be associated with the fracture risk [12] and bone loss [13] in the elderly, is used as an input variable for estimating the risk of fractures.

The flow of input variables and output information is shown in **Figure 1**. The web application’s input was designed using the web application concept, which does not store or save any input data to keep privacy and confidentiality for users. Additionally, BONEcheck allows users to create accounts to save their results for future comparison and also provides the option for users to delete their accounts if desired.

**Figure 1:**
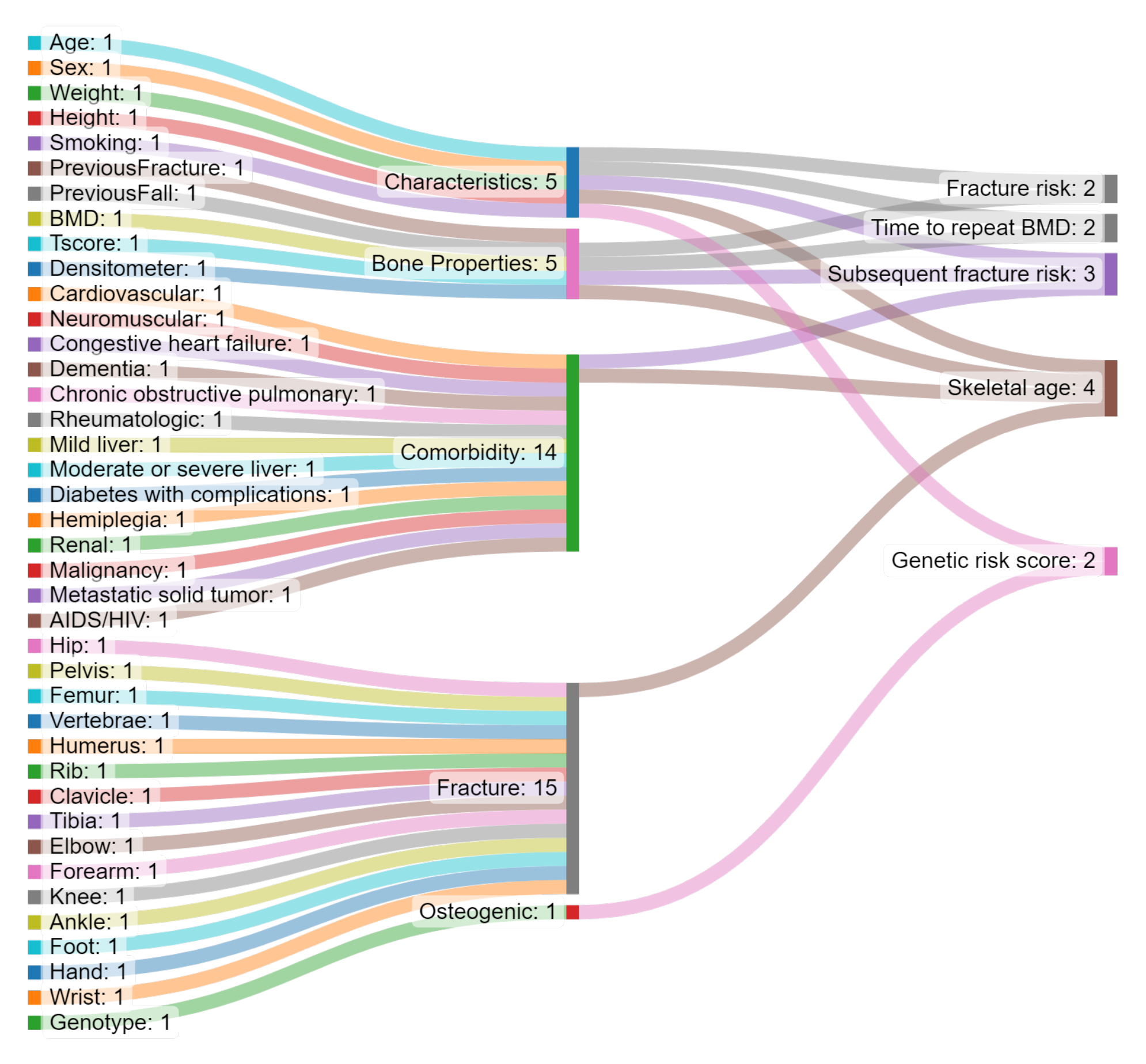
An example of output screen of BONEcheck.

### Output information

Based on the input variables, a series of output information is produced. These outputs are related to (a) the risk of fracture; (b) skeletal age; and (c) bone loss assessment. The risk of fracture is presented as the absolute probability of any fracture and hip fracture over the next 5 years. We chose to focus on 5-year window because that is the ideal time for an individual to manage their risk. In addition to numerical probability, we also provide a frequency of human icons to capture the risk visually. For instance, a 10% risk is presented as 10 red icons in 100 human icons. Because an existing fracture is a signal of further fractures, we also provide the risk of refracture if a fracture has been sustained. The risk of refracture is presented in a numerical probability over the next 2 years using the published model [14]. In addition, any fracture risk is classified as a risk gradient as follows: "high risk" if 5-year fracture probability exceeds 8%; "medium risk" if the probability ranges between 5 and 8%; and "low risk" if the probability is below 5%. When it comes to hip fracture, “high risk” signifies a risk greater than 2%, “medium risk” represents a risk between 1% and 2%, and “low risk” indicates a risk level lower than 1%.

*Skeletal age* is a new metric of fracture risk assessment. Conceptually, skeletal age is the age of an individual’s skeleton because of a fracture or being exposed to risk factors that elevate the risk of fracture [15]. Operationally, skeletal age is defined as the sum of an individual’s actual age and the years of life lost associated with a fracture or exposure to risk factors that put an individual at a greater risk of fracture. BONEcheck integrates the risk of fracture and the US lifetable to estimate the years of life lost associated with a fracture and then determined the skeletal age.

#### Bone loss assessment

Using published findings [16, 17], BONEcheck estimates the rate of change in femoral neck BMD for those aged 50 years and older stratified by baseline BMD. From the estimated rate of change, the algorithm uses linear regression to determine the time to reach ’osteoporosis’ (i.e., T-scores ≤ −2.5). There is additional advice that users need to consult with their doctors to determine the time to repeat BMD measurement.

### Interpretation/contextualization

In addition to risk estimates, BONEcheck provides interpretations of the probability of fracture tailored to an individual’s risk profile. The risk is presented in two scenarios: not treatment and on treatment. The interpretation is based on the ’frequentist’ school of probability, not subjective probability. Thus, a fracture probability of 10% is interpreted as 10 fractures in 100 men/women like the individual. The reduction of risk was derived from published results of randomized controlled trials [18]. The probability of fracture is then referred to the current Australian Pharmaceutical Benefits Scheme (PBS), which states that individuals with a 5-year fracture probability of 8-13% or higher are eligible for reimbursement.

For skeletal age, it explicitly acknowledges that a fracture is correlated with a reduction in life expectancy, with a corresponding interpretation provided. If an individual’s skeletal age surpasses their chronological age, it indicates that the individual is at a greater risk of fracture and mortality when compared to other people of the same age and gender. For those without a fracture, the number of years of life lost is determined as the product of the remaining lifetime risk of fracture and the number of years of life lost associated with a hip fracture, stratified by gender and age. Users are advised that by implementing preventative measures or following a bone specialist’s recommended effective treatment, they can decrease their skeletal age.

There is a ’Prevention’ tab where users can learn about preventive measures to reduce their risk of fracture and improve their BMD measurement. The advice given in the Prevention tab is based on current guidelines for the treatment and prevention of osteoporosis [19].

## Results

The web-based graphical interface of BONEcheck is shown in **Figure 2**. The tool collects each individual’s input data; processes the data; loads the algorithms or training models; calculates the metrics, and displays the results of calculation and interpretation. The risk of fracture is ’individualized’ in the sense that each individual has a unique probability of fracture which is calculated from the individual’s unique risk profile. This unique profile is defined in terms of the ’Osteogenomic profile’ and other clinical parameters. For illustration, **Table 2** presents the output of BONEcheck for 4 individuals:

- *Individual A*: woman, 65 years old, has sustained a hip fracture, 1 fall over the past 12 months, T-score is −2.0, has type 2 diabetes, no genetic profile data.
- *Individual B*: woman, 65 years old, has no fracture, no fall, T-score is −2.5, has congestive heart failure, genetic profile data.
- *Individual C*: man, 65 years old, has 1 vertebral fracture, 1 fall, T-score is −2.0, has type 2 diabetes, no genetic profile data.
- *Individual D*: man, 65 years old, has no vertebral fracture, no fall, T-score is −2.5, has COPD, genetic profile data.

**Figure 2:**
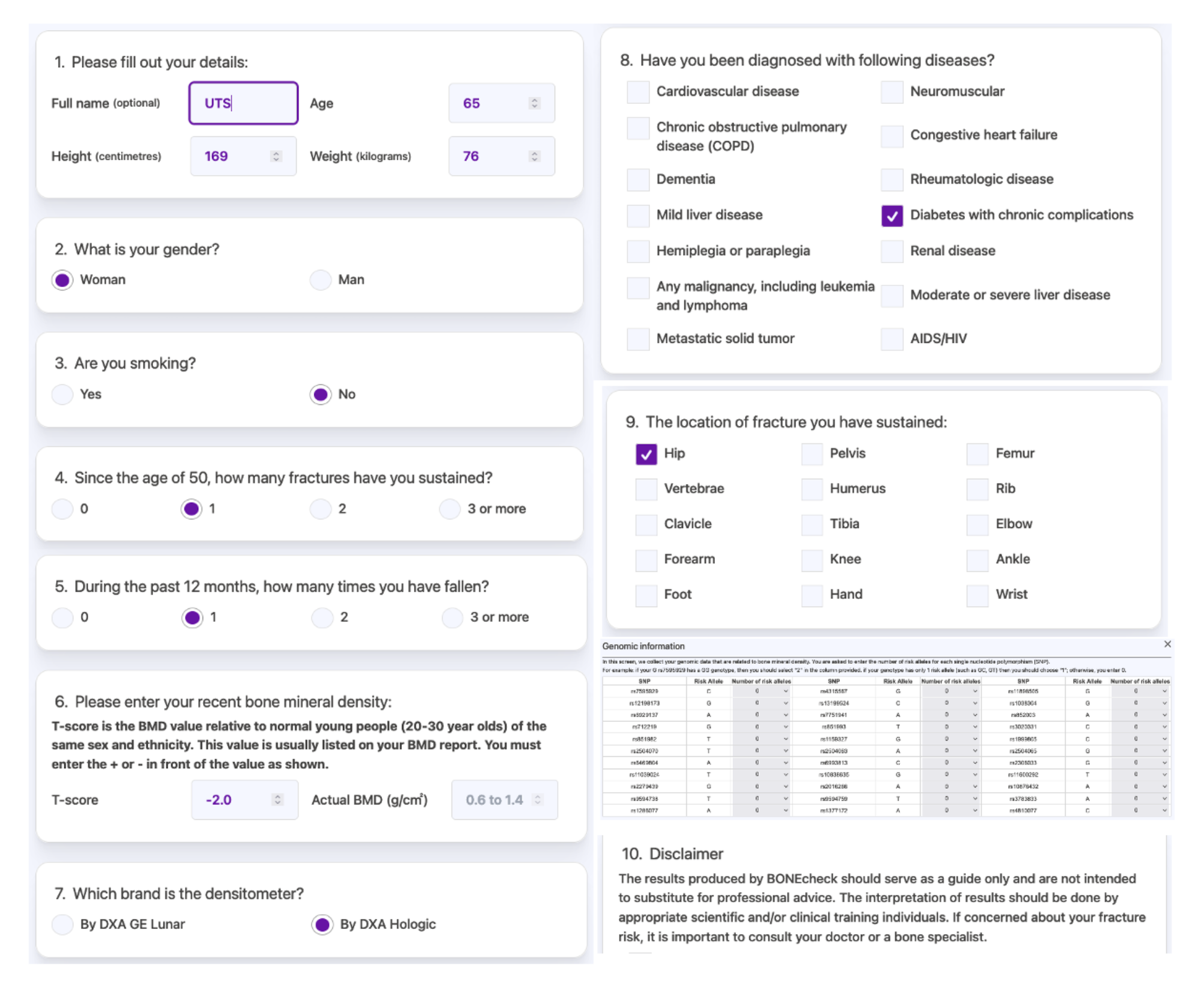
Input screen of BONEcheck.

**Table 2:** Illustration of BONEcheck with 4 hypothetical cases.

| **Profile** | **Individual**<br>**A** | **Individual**<br>**B** | **Individual**<br>**C** | **Individual**<br>**D** |
| --- | --- | --- | --- | --- |
| Gender | Woman | Woman | Man | Man |
| Age (years) | 65 | 65 | 65 | 65 |
| Prior fracture | Hip | No | Vertebral | No |
| Number of falls over the past 12 months | 1 | No | 1 | No |
| T-score | -2.0 | -2.5 | -2.0 | -2.5 |
| Comorbidity<sup>1-3</sup> | DM | CHF | DM | COPD |
| Genetic profile available | No | Yes | No | Yes |
| **Output** |  |  |  |  |
| 5-year risk of any fracture | 15% | 9% | 10% | 5% |
| 5-year risk of hip fracture | 5% | 2% | 20% | 1% |
| Risk of refracture | 30% | 12.6% | 2% | 8.1% |
| Skeletal age | 68.7 | 67.5 | 68.5 | 66.8 |
| Time to reach osteoporosis | 22 months |  | 20 months |  |

<sup>1</sup>DM: diabetes mellitus; <sup>2</sup>CHF: congestive heart failure; <sup>3</sup>COPD: chronic obstructive pulmonary disease

As can be seen from Table 2, for the same age and gender, the risk of fracture (any fracture and hip fracture) is inversely associated with femoral neck BMD T-scores, and for the same age and T-score, women, as expected, have a higher risk of fracture than men. For those with an existing fracture, the risk of refracture is relatively high for a shorter duration.

The risk of fracture is also presented in a human icon format (**Figure 3**). In this presentation of 100 icons, the ones with a fracture are shown in red colour, and the benefit of treatment (in terms of fracture risk reduction) is shown in green color. This is accompanied by an interpretation as follows (for individual A):

**Figure 3:**
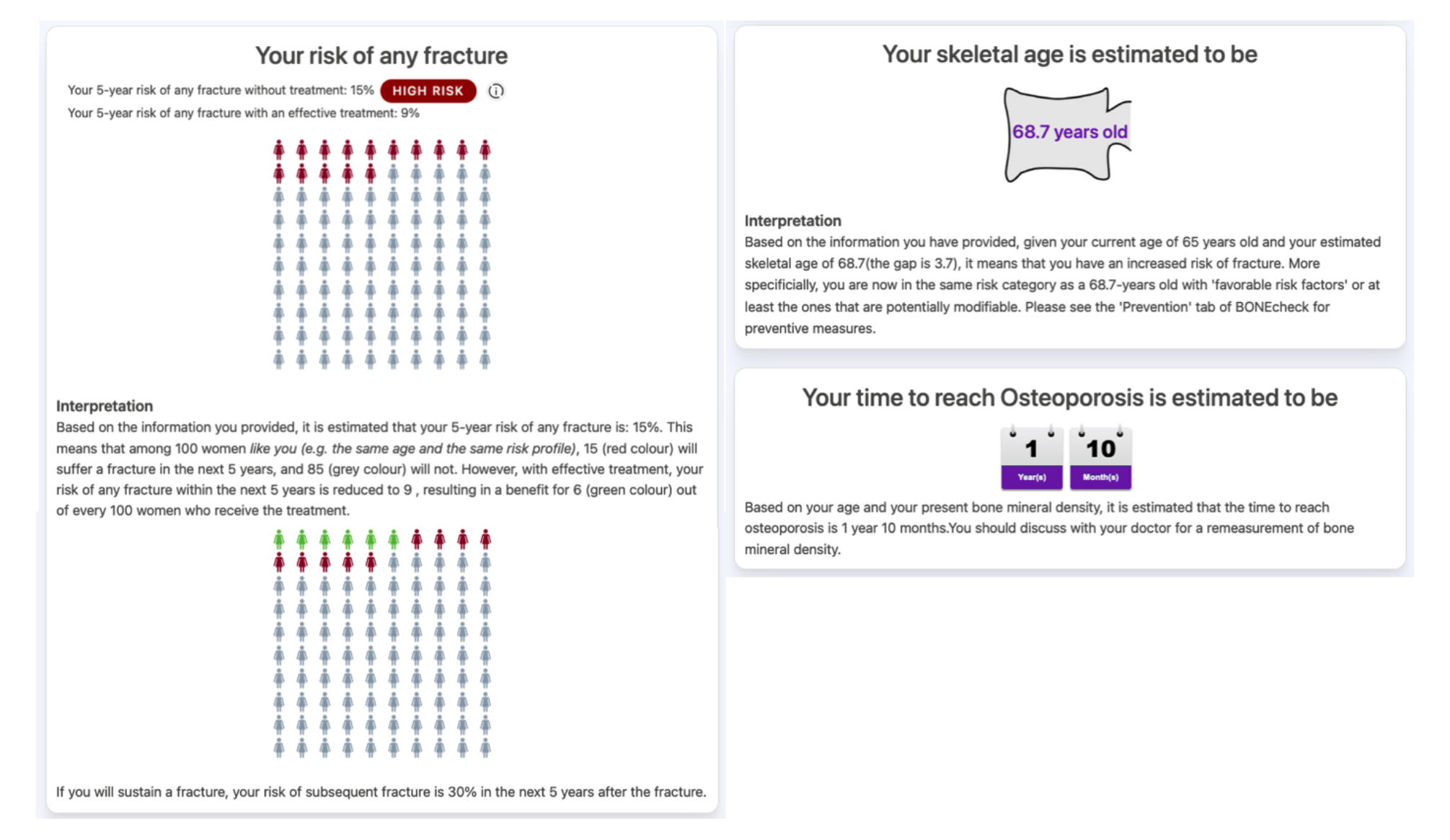
The flow of input variables and output in the BONEcheck system.

> *"Based on the information you provided, it is estimated that your 5-year risk of any fracture is: 15%. This means that among 100 women like you (e.g., the same age and the same risk profile), 15 (red colour) will suffer a fracture in the next 5 years, and 85 (grey colour) will not. However, with effective treatment, your risk of any fracture within the next 5 years is reduced to 9, resulting in a benefit for 6 (green colour) out of every 100 women who receive the treatment."*

A similar output and interpretation are also provided for hip fracture risk.

The skeletal age analysis shows that a hip fracture confers a greater impact on the years of life lost and results in a higher skeletal age than a vertebral fracture. A 65-year-old woman who has sustained a hip fracture is predicted to have a skeletal age of 68.3 years (individual A).

Moreover, the skeletal age of a 65-year-old man who has sustained a vertebral fracture is estimated to be 68.5 years (individual C). The skeletal age output is also accompanied by a graphical format (**Figure 3**) with an interpretation as follows (individual A):

> *"Based on the information you have provided, given your current age of 65 years old and your estimated skeletal age of 68.7(the gap is 3.7), it means that you have an increased risk of fracture. More specifically, you are now in the same risk category as a 68.7-years old with ’favorable risk factors’ or at least the ones that are potentially modifiable. Please see the ’Prevention’ tab of BONEcheck for preventive measures."*

The time to reach osteoporosis (i.e., T-scores ≤ −2.5) is estimated based on current age and BMD measurement. Thus, for an individual with a current T-score = −2.0, it is estimated that the time to reach osteoporosis (T < −2.5) is 22 months (for women) and 20 months (for men). If the current T-score ≤ −2.5, then the output will read "*You are having osteoporosis. You should discuss with your doctor for a treatment option and a repeat bone density measurement.*"

All output information can be saved into a file or an internet link so users can use it to discuss with their doctor. The saved information can also be used for longitudinal comparison for a user.

## Discussion

Fracture due to osteoporosis remains a significant burden worldwide because it is associated with an increased risk of premature mortality and substantial healthcare costs. An essential effort of fracture prevention focuses on the identification of high-risk individuals for intervention, and a number of risk assessment tools have been developed and implemented for this purpose.

However, none of the tools incorporates pre-mature mortality and risk contextualization. Moreover, the majority of osteoporosis patients are not treated or do not adhere to treatment guidelines due in part to poor risk communication. In order to address those shortcomings, we have developed the BONEcheck system for public use with the hope of contributing to the reduction of the global burden of osteoporosis.

The difference in output between BONEcheck and existing algorithms is shown in **Table 3**. Existing fracture risk assessment tools such as the Garvan Fracture Risk Calculator [6], FRAX [7], and Qfracture [20] provide 5-year [6] or 10-year [6, 7] risk of any fracture or osteoporotic fracture and hip fracture. The present BONEcheck utilizes the Garvan Fracture Risk Calculator algorithm to estimate 5-year risk of fracture. The predicted risk of fracture is different between algorithms due largely to underlying statistical models and input variables. Although FRAX’s predicted risk is adjusted to account for the competing risk of mortality, the details of how this adjustment was made have not been disclosed. On the other hand, the Garvan model was created using data from the Dubbo Osteoporosis Epidemiology Study, where each individual’s sequential events of fracture, refracture, and death were directly observed. As a result, the predicted risk inherently represents the likelihood of experiencing a fracture among those who are at risk for the remainder of their specific lifespan. In the Geelong Osteoporosis Study [21], the Garvan model underestimated fracture risk by around 25% in women and 19% in men, whereas FRAX underestimated it by 55% in women and 66% in men. In contrast, in the New Zealand cohort, the Garvan model predicted nearly 100% of fracture cases but overestimated hip fracture risk by 50%, while FRAX underestimated fracture risk by 50% [22]. However, the Garvan model’s overestimation has no negative clinical impact since high-risk individuals would be recommended for treatment regardless. There is evidence suggesting that the Garvan model’s predicted risk is consistent with the clinical decision-making [23, 24].

**Table 3:** Comparison between existing fracture risk assessment tools.

| **Tool functions** | **FRAX** | **Garvan** | **QFracture** | **BONEcheck** |
| --- | --- | --- | --- | --- |
| 5-year fracture risk | - | - | + | + |
| 10-year fracture risk | + | + | + | - |
| Subsequent fracture risk | - | - | - | + |
| Polygenic risk score | - | - | - | + |
| Skeletal age | - | - | - | + |
| Time to reach osteoporosis | - | - | - | + |
| Risk contextualization | - | - | - | + |
| Interpretation | - | - | + | + |

At present, the management of osteoporosis is in a crisis of an ‘osteoporosis treatment gap’ characterized by low treatment uptake among those at high risk. Hip fracture is the most serious consequence of osteoporosis because patients suffering from the fracture are at increased risk of mortality. There are treatments that have been shown to reduce the risk of refractures and mortality [25, 26]. However, few patients with hip fractures were on treatment. In 2001, 40% of hip fracture patients were on treatment, and this proportion decreased to only 21 in 2011 [5].

Moreover, among those on treatment, adherence has been poor [27]. There are many reasons for the treatment gap, including the problem of risk communication. According to a qualitative study, patients who believed that osteoporosis was a natural part of aging, that treatment was ineffective, and that fractures were not serious chose not to pursue or discontinue treatment [28-30]. Moreover, patients were more likely to accept treatment if they were presented with the benefit of treatment in terms of absolute risk reduction rather than relative risk reduction [31]. Taken together, the research evidence indicates the necessity for innovative approaches to risk communication in order to enhance the adoption and compliance with anti-osteoporosis treatment.

Currently, patients are presented with fracture probabilities over a specific time frame without any quantitative information about the consequences of a fracture. Additionally, there is no clear indication of the survival advantages of treatment. To address these issues, BONEcheck was developed. BONEcheck not only offers absolute fracture risk assessments but also frames the absolute risk reduction and the survival benefits of treatment. This information can be valuable for facilitating productive discussions between doctors and patients regarding risk, treatment, and benefits. The risk information and treatment benefits are shown in the human icon array rather than numerical format [32].

In addition to offering a predicted risk of fracture, BONEcheck capitalizes on recent research to provide information on the skeletal age [15]. Skeletal age is a way to understand the risk of fracture and its consequence of premature mortality. An individual’s risk of fracture may be higher if the individual’s skeletal age is greater than their chronological age. In this context, if an individual is 60 years old but has a skeletal age of 65, it implies that the individual’s fracture risk matches that of a 65-year-old person with a ’healthy’ risk profile or at least ones that can be modified. Previous studies suggested that conveying risk using biological age indicators, such as heart age, vascular age, lung age, and skeletal age, could potentially have a favorable influence on patient behavior [33]. Since most fractures occur outside the high-risk (osteoporosis) group [34], risk communication using skeletal age (’older than I actually am’) can help raise awareness of the mortality consequence of fracture in those groups.

In addition to risk communication (e.g., risk probability and skeletal age), BONEcheck also presents data on the estimated duration it would take for individuals who are currently not classified as having osteoporosis to develop the condition. This information can be especially useful for facilitating discussions between doctors and patients regarding the appropriate timeframe for repeating bone mineral density measurements.

BONEcheck is the first tool that incorporates the polygenic risk score (PRS) to predict fracture risk. Several PRSes have been formulated [35-37] based on the identification of genetic variants linked to BMD or fracture, in addition to our own [12]. These PRSes utilize different genetic variants, but each has been validated as an independent predictor of fracture risk beyond clinical risk factors. Our PRS, which was created from 33 genetic variants associated with BMD, can replace family history as a fracture risk factor. Assessing the likelihood of fracture, or any disease risk, should be personalized because no "average individual" exists in the population, and each person is unique. An individual’s distinctiveness can be characterized in terms of clinical risk factors, as well as PRS.

The algorithms used to create BONEcheck were derived from data obtained from Caucasian populations which may have a higher fracture risk compared to Asian populations. Thus, the extrapolation of risk from BONEcheck to non-Caucasian requires certain adjustments. Furthermore, there is a lack of evidence from randomized controlled trials indicating that intervening in individuals at high risk of fracture results in a decrease in fracture risk. Despite this, the output provided by BONEcheck can be beneficial for promoting discussions between doctors and patients regarding the prevention of osteoporotic fractures.

In conclusion, we have developed and implemented a digital tool called ’BONEcheck’ for fracture risk assessment which is now available free worldwide. The tool can help facilitate doctor-patient discussion about fracture risk, clinical consequences, and treatment benefits so that an informed decision can be reached.

## Data Availability

All data produced in the present study are available upon reasonable request to the authors

## Acknowledgments

The development of BONEcheck is supported in part by a grant from the National Health and Medical Research Council in Australia (APP1195305) and Amgen Competitive Grant Program (2019). Neither the funding sources nor the authors’ institutions had any role in the study design; in the collection, analysis and interpretation of data; in the writing of the report; and in the decision to submit this manuscript for publication.

## Availability

BONEcheck is now accessible to users through multiple platforms. Users can access it directly from our website or download the app from the Apple Store or Google Play. Please click on the links below to start utilizing the BONEcheck tool:

Website: https://bonecheck.org/

Apple Store: https://apps.apple.com/app/bonecheck/id6447424513

Google Play: https://play.google.com/store/apps/details?id=org.saigonmec.bonecheck

Auto access: https://onelink.to/8cjb7m

QR code:

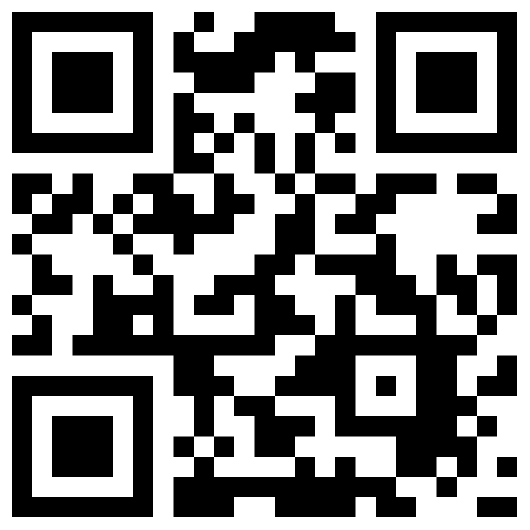

## References

1. Sozen T, Ozisik L, Basaran NC. An overview and management of osteoporosis. Eur J Rheumatol. 2017;4(1):46–56. Epub 2017/03/16. doi: 10.5152/eurjrheum.2016.048. PubMed PMID: 28293453; PubMed Central PMCID: PMCPMC5335887.

2. Collaborators GBDF. Global, regional, and national burden of bone fractures in 204 countries and territories, 1990-2019: a systematic analysis from the Global Burden of Disease Study 2019. Lancet Healthy Longev. 2021;2(9):e580–e92. doi: 10.1016/S2666-7568(21)00172-0. PubMed PMID: 34723233; PubMed Central PMCID: PMCPMC8547262.

3. Center JR, Bliuc D, Nguyen TV, Eisman JA. Risk of subsequent fracture after low-trauma fracture in men and women. Jama. 2007;297(4):387–94. PubMed PMID: 17244835.

4. Bliuc D, Nguyen ND, Milch VE, Nguyen TV, Eisman JA, Center JR. Mortality risk associated with low-trauma osteoporotic fracture and subsequent fracture in men and women. Jama. 2009;301(5):513–21. Epub 2009/02/05. doi: 301/5/513 [pii] 10.1001/jama.2009.50. PubMed PMID: 19190316.

5. Miller PD. Underdiagnosis and Undertreatment of Osteoporosis: The Battle to Be Won. J Clin Endocrinol Metab. 2016;101(3):852–9. doi: 10.1210/jc.2015-3156. PubMed PMID: 26909798.

6. Nguyen ND, Frost SA, Center JR, Eisman JA, Nguyen TV. Development of a nomogram for individualizing hip fracture risk in men and women. Osteoporos Int. 2007;18(8):1109–17. Epub 2007/03/21. doi: 10.1007/s00198-007-0362-8. PubMed PMID: 17370100.

7. Kanis JA, Johnell O, Oden A, Johansson H, McCloskey E. FRAX and the assessment of fracture probability in men and women from the UK. Osteoporos Int. 2008;19(4):385–97. Epub 2008/02/23. doi: 10.1007/s00198-007-0543-5. PubMed PMID: 18292978; PubMed Central PMCID: PMC2267485.

8. Trevena LJ, Zikmund-Fisher BJ, Edwards A, Gaissmaier W, Galesic M, Han PK, et al. Presenting quantitative information about decision outcomes: a risk communication primer for patient decision aid developers. BMC Med Inform Decis Mak. 2013;13 Suppl 2(Suppl 2):S7. Epub 20131129. doi: 10.1186/1472-6947-13-S2-S7. PubMed PMID: 24625237; PubMed Central PMCID: PMCPMC4045391.

9. Gigerenzer G, Gaissmaier W, Kurz-Milcke E, Schwartz LM, Woloshin S. Helping Doctors and Patients Make Sense of Health Statistics. Psychol Sci Public Interest. 2007;8(2):53–96. Epub 20071101. doi: 10.1111/j.1539-6053.2008.00033.x. PubMed PMID: 26161749.

10. Charlson ME, Pompei P, Ales KL, MacKenzie CR. A new method of classifying prognostic comorbidity in longitudinal studies: development and validation. J Chronic Dis. 1987;40(5):373–83. Epub 1987/01/01. doi: 10.1016/0021-9681(87)90171-8. PubMed PMID: 3558716.

11. Styrkarsdottir U, Halldorsson BV, Gretarsdottir S, Gudbjartsson DF, Walters GB, Ingvarsson T, et al. Multiple genetic loci for bone mineral density and fractures. N Engl J Med. 2008;358(22):2355–65. Epub 2008/05/01. doi: NEJMoa0801197 [pii] 10.1056/NEJMoa0801197. PubMed PMID: 18445777.

12. Ho-Le TP, Center JR, Eisman JA, Nguyen HT, Nguyen TV. Prediction of Bone Mineral Density and Fragility Fracture by Genetic Profiling. J Bone Miner Res. 2017;32(2):285–93. doi: 10.1002/jbmr.2998. PubMed PMID: 27649491.

13. Ho-Le TP, Pham HM, Center JR, Eisman JA, Nguyen HT, Nguyen TV. Prediction of changes in bone mineral density in the elderly: contribution of "osteogenomic profile". Archives of osteoporosis. 2018;13(1):68. doi: 10.1007/s11657-018-0480-2. PubMed PMID: 29931598.

14. Ho-Le TP, Tran TS, Bliuc D, Pham HM, Frost SA, Center JR, et al. Epidemiological transition to mortality and refracture following an initial fracture. Elife. 2021;10. Epub 2021/02/10. doi: 10.7554/eLife.61142. PubMed PMID: 33558009; PubMed Central PMCID: PMCPMC7924952.

15. Tran TS, Ho-Le TP, Bliuc D, Abrahamsen B, Hansen L, Vestergaard P, et al. ‘Skeletal Age’ for mapping the impact of fracture on mortality. eLife. 2023; In-press.

16. Nguyen TV, Center JR, Eisman JA. Femoral neck bone loss predicts fracture risk independent of baseline BMD. J Bone Miner Res. 2005;20(7):1195–201. doi: 10.1359/JBMR.050215. PubMed PMID: 15940372.

17. Ensrud KE, Palermo L, Black DM, Cauley J, Jergas M, Orwoll ES, et al. Hip and calcaneal bone loss increase with advancing age: longitudinal results from the study of osteoporotic fractures. J Bone Miner Res. 1995;10(11):1778–87. PubMed PMID: Department of Medicine, VA Medical Center, Minneapolis, Minnesota, USA.

18. Cummings SR, San Martin J, McClung MR, Siris ES, Eastell R, Reid IR, et al. Denosumab for prevention of fractures in postmenopausal women with osteoporosis. N Engl J Med. 2009;361(8):756–65. Epub 2009/08/13. doi: 10.1056/NEJMoa0809493. PubMed PMID: 19671655.

19. Khosla S, Wright NC, Elderkin AL, Kiel DP. Osteoporosis in the USA: prevention and unmet needs. Lancet Diabetes Endocrinol. 2023;11(1):19–20. Epub 20221114. doi: 10.1016/S2213-8587(22)00322-9. PubMed PMID: 36395774.

20. Hippisley-Cox J, Coupland C. Derivation and validation of updated QFracture algorithm to predict risk of osteoporotic fracture in primary care in the United Kingdom: prospective open cohort study. BMJ. 2012;344:e3427. doi: 10.1136/bmj.e3427. PubMed PMID: 22619194.

21. Holloway-Kew KL, Zhang Y, Betson AG, Anderson KB, Hans D, Hyde NK, et al. How well do the FRAX (Australia) and Garvan calculators predict incident fractures? Data from the Geelong Osteoporosis Study. Osteoporos Int. 2019;30(10):2129–39. Epub 2019/07/19. doi: 10.1007/s00198-019-05088-2. PubMed PMID: 31317250.

22. Bolland MJ, Siu AT, Mason BH, Horne AM, Ames RW, Grey AB, et al. Evaluation of the FRAX and Garvan fracture risk calculators in older women. J Bone Miner Res. 2011;26(2):420–7. Epub 2010/08/20. doi: 10.1002/jbmr.215. PubMed PMID: 20721930.

23. Inderjeeth CA, Raymond WD. Case finding with GARVAN fracture risk calculator in primary prevention of fragility fractures in older people. Arch Gerontol Geriatr. 2020;86:103940. Epub 2019/09/17. doi: 10.1016/j.archger.2019.103940. PubMed PMID: 31525558.

24. Stuckey B, Magraith K, Opie N, Zhu K. Fracture risk prediction and the decision to treat low bone density. Aust J Gen Pract. 2021;50(3):165–70. Epub 2021/02/27. doi: 10.31128/AJGP-04-20-5363. PubMed PMID: 33634286.

25. Lyles KW, Colon-Emeric CS, Magaziner JS, Adachi JD, Pieper CF, Mautalen C, et al. Zoledronic acid and clinical fractures and mortality after hip fracture. N Engl J Med. 2007;357(18):1799–809. PubMed PMID: 17878149.

26. Reid IR, Horne AM, Mihov B, Stewart A, Garratt E, Bastin S, et al. Effects of Zoledronate on Cancer, Cardiac Events, and Mortality in Osteopenic Older Women. J Bone Miner Res. 2020;35(1):20–7. Epub 2019/10/12. doi: 10.1002/jbmr.3860. PubMed PMID: 31603996.

27. Fatoye F, Smith P, Gebrye T, Yeowell G. Real-world persistence and adherence with oral bisphosphonates for osteoporosis: a systematic review. BMJ open. 2019;9(4):e027049. Epub 2019/04/17. doi: 10.1136/bmjopen-2018-027049. PubMed PMID: 30987990; PubMed Central PMCID: PMCPMC6500256.

28. Salter C, Brainard J, McDaid L, Loke Y. Challenges and opportunities: what can we learn from patients living with chronic musculoskeletal conditions, health professionals and carers about the concept of health literacy using qualitative methods of inquiry? PLoS One. 2014;9(11):e112041. Epub 2014/11/07. doi: 10.1371/journal.pone.0112041. PubMed PMID: 25375767; PubMed Central PMCID: PMCPMC4222964.

29. Hawarden A, Jinks C, Mahmood W, Bullock L, Blackburn S, Gwilym S, et al. Public priorities for osteoporosis and fracture research: results from a focus group study. Archives of osteoporosis. 2020;15(1):89. Epub 2020/06/18. doi: 10.1007/s11657-020-00766-9. PubMed PMID: 32548718; PubMed Central PMCID: PMCPMC7297850.

30. Paskins Z, Crawford-Manning F, Cottrell E, Corp N, Wright J, Jinks C, et al. Acceptability of bisphosphonates among patients, clinicians and managers: a systematic review and framework synthesis. BMJ open. 2020;10(11):e040634. Epub 2020/11/06. doi: 10.1136/bmjopen-2020-040634. PubMed PMID: 33148763; PubMed Central PMCID: PMCPMC7640526.

31. Ralston KAP, Phillips J, Krause A, Hauser B, Ralston SH. Communicating Absolute Fracture Risk Reduction and the Acceptance of Treatment for Osteoporosis. Calcif Tissue Int. 2022;110(6):698–702. Epub 2022/02/14. doi: 10.1007/s00223-022-00948-2. PubMed PMID: 35152304; PubMed Central PMCID: PMCPMC9108104.

32. Elwyn G, Price A, Franco JVA, Gulbrandsen P. The limits of shared decision making. BMJ Evid Based Med. 2022. Epub 2022/12/16. doi: 10.1136/bmjebm-2022-112089. PubMed PMID: 36522136.

33. Kulendrarajah B, Grey A, Nunan D. How effective are ’age’ tools at changing patient behaviour? A rapid review. BMJ Evid Based Med. 2020;25(2):1–2. Epub 2019/09/29. doi: 10.1136/bmjebm-2019-111244. PubMed PMID: 31558486.

34. Mai HT, Tran TS, Ho-Le TP, Center JR, Eisman JA, Nguyen TV. Two-thirds of all fractures are not attributable to osteoporosis and advancing age: implication for fracture prevention. J Clin Endocrinol Metab. 2019. doi: 10.1210/jc.2018-02614. PubMed PMID: 30951170.

35. Lee SH, Cho EH, Ahn SH, Kim HM, Lim KH, Kim BJ, et al. Prediction of Future Osteoporotic Fracture Occurrence by Genetic Profiling: A 6-Year Follow-Up Observational Study. J Clin Endocrinol Metab. 2016;101(3):1215–24. Epub 20160112. doi: 10.1210/jc.2015-3972. PubMed PMID: 26756118.

36. Eriksson J, Evans DS, Nielson CM, Shen J, Srikanth P, Hochberg M, et al. Limited clinical utility of a genetic risk score for the prediction of fracture risk in elderly subjects. J Bone Miner Res. 2015;30(1):184–94. doi: 10.1002/jbmr.2314. PubMed PMID: 25043339; PubMed Central PMCID: PMCPMC4281709.

37. Lu T, Forgetta V, Keller-Baruch J, Nethander M, Bennett D, Forest M, et al. Improved prediction of fracture risk leveraging a genome-wide polygenic risk score. Genome Med. 2021;13(1):16. Epub 20210203. doi: 10.1186/s13073-021-00838-6. PubMed PMID: 33536041; PubMed Central PMCID: PMCPMC7860212.

